# Medical Abortion Self Use in Kenya: Results from a process evaluation of women’s experiences

**DOI:** 10.1101/2022.11.10.22282174

**Authors:** Japheth O. Ouma, Linah A. Musimbi, Edward O. Ngoga, Steve B. Sigu, Beatrice A. Otieno, Angela Akol

**Affiliations:** Ipas Africa Alliance, P.O. Box 1192-00200 City Square, Nairobi-Kenya

## Abstract

**Background:** The consequences of unsafe abortions weigh heavily on individuals, families, society, and health care systems. Developing countries continue to experience a high prevalence of mistimed and unintended pregnancies resulting in induced abortions that are largely unsafe. Globally, 25 million unsafe abortions take place every year [1]. In Africa, 3 out of 4 abortions are categorized as unsafe [2]. In Kenya, it is estimated that 464,690 induced abortions occur annually- 2 out of 3 of these are unsafe[3]. With the high prevalence of mistimed and unwanted pregnancies in Sub-Saharan Africa and Kenya in particular, unsafe abortions will be on the rise unless deliberate measures and policies are put in place to guarantee safe abortion procedures[4].

**Method:** Ipas Africa Alliance embarked on the Medical Abortion Self-Use (MASU) project aimed at reducing morbidity and mortality tied to unintended pregnancies among women in the Counties of Vihiga, Kisumu, Busia, Siaya, and Trans Nzoia. To inform on the progress and the potential for scale-up of the project, Ipas commissioned a process evaluation. This evaluation adopted both qualitative and quantitative methods. This involved in-depth interviews (IDIs), Focus Group Discussions (FGDs), and face-face-interviews through a semi-structured questionnaire.

**Results:** From the analysis, those seeking medical abortion were mainly youths under 25 years of age. There was low awareness of safe abortion practices and the gestation period within which safe medical abortion (MA) can be safely done. Nearly half (47%) of the women and girls reported being coerced to take post-abortion contraceptives they never wanted. Further, MA costs were not only found to be expensive but also varied greatly across the Counties. Some MA users experienced medical complications attributed to the failure of pharmacists responsible to provide the correct dose and correct route of administration. On the other hand, youth champions were found to be few and not able to fully cover their areas and only have basic knowledge of MA self-use services.

**Conclusion:** The IPAS MASU project intervention, in the five counties of Western Kenya namely Busia, Siaya, Vihiga, Kisumu, and Trans Nzoia, has increased access to safe Medical Abortion self-use, enhanced availability of MA drugs in pharmacies, the improved service delivery of MA services through regular training of pharmacists. Further, the project has enhanced awareness about MA services among young girls and women through trained Youth Champions and pharmacists, and the MASU project has significantly reduced cases of unsafe abortions and by extension deaths and medical complications associated with them. To realize more gains, the project needs to be scaled up within the five counties and beyond, specifically to target the rural areas where cases of unsafe abortion are still thought to be rampant.

## 1.0 BACKGROUND

Developing countries continue to experience a high prevalence of mistimed and unintended pregnancies resulting in induced abortions that are largely unsafe. Globally, 25 million unsafe abortions take place every year [1]. [1]. In Africa, 3 out of 4 abortions are categorized as unsafe [2]. In Kenya, it is estimated that 464,690 induced abortions occur annually-2 out of 3 of these are unsafe[3]. The consequences of unsafe abortions thus weigh heavily on individuals, families, society, and the health care systems. In Kenya, maternal deaths directly associated with unsafe abortion are nearly 6,000 yearly [5]. Out of these maternal deaths, about 5000 occur in low-income areas and rural set-ups [5].

Though the World Health Organization (WHO) developed policy guidelines on safe abortions for pregnancies up to 9 weeks gestation (63 days)[6], medical abortion (MA) self-use is yet to be fully operationalized in developing countries. The western part of Kenya (Counties of Nyanza and Western) is the highest after Rift Valley in incidences of unsafe abortions[7]. The issue is compounded by the low awareness about safe abortion practices, especially in the rural areas of these counties [8]. Against this backdrop, Ipas initiated the MASU project aimed at deepening access to information on sexual reproductive health and rights and reducing morbidity and mortality tied to unintended pregnancies among women in the Counties of Vihiga, Kisumu, Siaya, Busia, and Trans Nzoia by deepening the use of medical abortion (MA).

IPAS works globally to improve access to safe abortion and contraception so that every woman and girl can be able to make informed reproductive decisions. Across Kenya, the organization works with partners (Ministry of Health, pharmacists, MA suppliers) to make safe abortion and contraception widely accessible to women and girls. To inform on the scale-up of the MASU project and monitor how well the project plans and activities are working and hence, guide interventions adjustments on account of any problems detected, Ipas carried out process evaluation in the five counties (Vihiga, Kisumu, Siaya, Busia, and Trans Nzoia) where the project currently runs. The evaluation gathered information on the awareness of MA and the MASU project, access, and utilization of MA (safe abortions), and post-MA contraception use that can be attributed to the interventions by the MASU project in the targeted and proximal counties.

## 2.0 METHODOLOGY

### 2.1 Evaluation Design

To achieve the stated objectives, the process evaluation adopted a mixed-method approach in which qualitative information was used to complement quantitative findings. This consisted of quantitative (survey), and qualitative (In-depth interviews, Key Informant Interviews, and Focus Group Discussions). The use of the mixed method in the study design was informed by the fact that through the mixed methods, a researcher can gather the information that guarantees a deeper understanding of the issues under investigation and also gives a voice to the participants, enables generalizations of the study results to the general population, and also makes it possible to triangulate and complement both the quantitative and qualitative results [9, 10].

To contextualize the study, comprehensive and analytical desk reviews were also conducted. The desktop review analyzed Ipas MASU project documents that included MLE Log Frame, Expanding Medical Abortion Self-Use in East Africa, MASU Mapping Findings, Self-Management of Medical Abortion survey data, and project technical report for activities conducted during the project implementation phase.

### 2.2 Ethics approval

The study protocol was reviewed and approved by the Jaramogi Oginga Odinga Teaching and Referral Hospital (JOOTRH) Ethics and Scientific Research Committee, approval number: IERC/JOOTRH/464/21. Additional permits to conduct the study were obtained from the registered pharmacies’ proprietors and individual written consent was obtained from each FGD participant. During the informed consent process, data collectors emphasized to the participants that the study is voluntary and ensured that the participants understood the risks and benefits associated with the study and that they were ready and willing to participate. We do hereby confirm that all methods were carried out per relevant guidelines and regulations within the approved protocol.

### 2.3 Study Sample

Respondents consisted of the youth champions, pharmacists, MA drug suppliers, Ipas staff (directly involved in the implementation of the MASU Project), and women and girls (users who had benefited from the MASU project).

The youth (peer) champions are individuals contracted by Ipas to provide demand-creation activities including disseminating information and providing escorted referrals to women and girls in need of PAC/MA/post-abortion contraception services from Ipas-affiliated pharmacists. The providers and pharmacists act as centers where access to safe abortion self-care services and post-medical abortion contraception are offered to women and girls who had been given MA services. The drug suppliers provided the MA drugs on a demand basis by the pharmacists thereby, ensuring, there is no shortage of the needed drugs by the users. The users are the women and girls requiring PAC/MA/post-abortion contraception services. The Ipas staff were interviewed to gain their perspectives on the bottlenecks encountered in the implementation of the MASU Project, measures adopted to address the existing gaps and to provide information on the estimated project budget for reference in case of project scale-up.

#### 2.3.1 Inclusion and Exclusion criteria

Pharmacists with at least a monthly caseload of 12 MA clients were selected for the Key Informant Interviews. Given the sensitive nature of abortion, women and girls who participated in the study were purposively identified and recruited for the study-only those who had sought MA services from the provider’s pharmacists under the MASU project were included. Those who had not sought these services from the MASU project provider’s pharmacists were excluded. Peer champions who were in the counties of the study were purposively sampled, those in other areas were excluded.

Table 1 presents a summary of the sampling distributions used to provide information for this process evaluation.

**Table 1:** presents a summary of the sampling distributions.

| County | Survey (MA Users) | FGD (MA Users) | IDIs | KII |
| --- | --- | --- | --- | --- |
| Vihiga | 25 | 8 | 4 | 3 |
| Trans Nzoia | 17 | 7 | 2 | 4 |
| Siaya | 17 | 8 | 2 | 3 |
| Kisumu | 17 | 8 | 1 | 4 |
| Busia | 12 | 7 | 1 | 2 |
| Ipas Staff |  |  |  | 1 |
| **Total** | **88** | **38** | **10** | **17** |

### 2.4 Data Collection

Robust methods for the collection of both quantitative and qualitative data were employed in the study. These included face-to-face interviews, In-depth interviews (IDIs), Key informant Interviews (KII), and focus group Discussions (FGDs). The face-to-face interviews and focus group Discussions (FGDs) targeted the users of the MA services, women and girls in the 5 counties. On the other hand, In-depth interviews (IDIs) targeted selected users. The KII was administered to selected pharmacists in each of the five counties, youth champions, a representative of the IPAS MASU program team, and suppliers of medical abortion medicines.

A total of 88 women and users of MA service were interviewed by trained research assistants using Mobile Application and Reporting System (MARS) through the Open Data Kit (ODK) loaded in mobile devices. The data sets were then downloaded and analyzed, and a report was produced.

For the FGDs, a team of 8 selected women and girls who had used the MA services were assembled in halls with the consultant and trained research assistants who then guided the discussions and or interviews from a set of well-designed questions. Responses were then recorded and captured in short notes after each set of questions was read and the interviewer only proceeded to the next question whenever the team had exhausted their thoughts on a particular question.

### 2.5 Data Analysis

Quantitative data was downloaded from MARS in an Excel format. The data was cleaned, organized, and uploaded to Stata version 15 for analysis. Descriptive and regression analyses were then conducted. The results were then triangulated by information from the analyzed qualitative data. The results are presented in terms of tables, pie charts, and graphs.

## 3.0 EVALUATION FINDINGS

### 3.1 Introduction

Data collected for this process evaluation contains information delineated in terms of demographic characteristics, socio-economic features, and those aligned to supply and demand when it comes to Sexual and Reproductive Health issues including Medication Abortion. This section presents a discussion on key variables for demographic and socio-economic characteristics, and their association with the programmatic outcomes and delves deeper into the supply and demand issues on Sexual and Reproductive Health.

### 3.2 Respondent’s Sociodemographic characteristics

The majority of the survey respondents were in the age bracket of 20-24; 52 (60%). This was followed by those who were 25 years and above 26 (30%). Almost half of the respondents 40 (46%) reported that they were students. Table 2 presents a summary of the socioeconomic and demographic characteristics of the respondents.

**Table 2:** Socioeconomic and demographic characteristics of the respondents

| Indicator | Frequency (n) | Percent (%) |
| --- | --- | --- |
| **Age** |  |  |
| < 20 years | 9 | 10 |
| 20-24 years | 52 | 60 |
| 25 and above years | 26 | 30 |
| **Age at first birth** |  |  |
| 13-18 years | 14 | 34 |
| 19-24 years | 23 | 56 |
| 25-28 years | 4 | 10 |
| **Sources of Income** |  |  |
| Unemployed/not working | 16 | 9 |
| Student | 40 | 46 |
| Self-informal (business, farmer) | 24 | 27 |
| Formal employment | 8 | 18 |
| **Respondents' Highest Level of Education** |  |  |
| No education level | 15 | 17 |
| Completed Primary | 9 | 10 |
| Completed Secondary | 36 | 41 |
| Completed tertiary college | 23 | 26 |
| Completed University | 5 | 6 |
| **Marital Status** |  |  |
| Married | 73 | 83 |
| Single | 12 | 14 |
| Widow | 1 | 1 |
| Divorced | 1 | 1 |
| Separated | 1 | 1 |

Further analysis of the variable age has revealed that the mean age of the respondents was 20.6 (nearly 21 years). On the other hand, the median age stood at 20.5 years. The youngest respondent was 13 years old, while the oldest was 28 years of age.

### 3.3 Medical Abortion Services

#### 3.3.1 Awareness of Safe Abortion Services

The majority of the respondents 81 (92%) reported that they were not aware of safe abortion practices at the discovery of pregnancy, with only 7 (8%) of the respondents being informed of safe abortion access points. The same views were confirmed by a majority of the FGD participants and the IDs respondents. During the FGDs and IDIs, the respondents stated various sources of safe abortion information as summarized in text box 1.

##### Text Box 1

**Views of FGD Participants (All the 5 counties) on the Sources of Information for safe abortion services**.

###### Friends and relatives

two respondents stated that they got information from their close friends and a cousin to one of the respondents.

###### Google and pharmacist

one respondent said when she sought abortion options, she Googled for information and further inquired more from the pharmacy and procured the services after she felt satisfied.

###### Pharmacy

Two others confirmed to have visited pharmacy for information.

###### Social media

all the respondents shared that with various social media sites and platforms, the information on MASU is readily available, one shared that she got information from IPAs platforms.

On the issue of the adequacy of the availability of MA self-use services in each of the 5 counties, the majority of the respondents (Users, youth champions, and pharmacists) were in agreement that the IPAS MASU project is yet to serve their respective counties adequately, more so the rural areas, as the demand for the MA services is growing steadily. The specific views of the MA users from different counties are presented in text box 2.

##### Text Box 2

**IDI and KII Participants on the adequacy of the MA services due to the MASU Project**.

###### R (IDI-Siaya)

“No, I feel it’s not adequately served. There are more pharmacies that have not accessed their services.”

###### R (KII-Busia)

“I can talk of my place, which I feel is not adequate and more needs to be done. Am sure, the cases I handle really don’t reflect the situation across the entire county, especially the rural areas, meaning that if something more is done, we will get more cases coming to seek the services.”

###### R (IDI-Kisumu)

“…they should increase their reach in the country. In the county I don’t know how many are on board, but if they can reach out to more.”

###### R (IDI-Trans Nzoia)

“IPAS should reach somewhere and try to help us in terms of contraceptives. There was a time we had shortages of contraceptives, especially the three months injection…”

###### R (KII-Suppliers 1&2)

There has been a substantial rise in demand for medical abortion self-use drugs in the counties of intervention compared with the rest.

###### R (KII-Suppliers 2)

Yes, we meet the current market demand

#### 3.3.2 Where Medical Abortion Services were Obtained

Respondents mentioned pharmacies and chemists 67 (76%) as the leading and most convenient access points owing to guaranteed privacy and friendly service delivery. This was followed by clinics 8 (9%) and private hospitals 4 (5%). Home as a place of abortion was the least mentioned. This is presented in Table 3.

**Table 3:** Place where Abortion was done.

| Place | Frequency (n) | Percent (%) |
| --- | --- | --- |
| Pharmacy/Chemists | 67 | 76% |
| Clinic | 8 | 9% |
| Public Hospital | 2 | 2% |
| Private Hospital | 4 | 5% |
| Others (Specify) | 7 | 8% |

The results in Table 4, identifying pharmacies as the most preferred place, by girls and women, for safe MA self-use services were corroborated by pharmacists from Busia, Kisumu, and Trans Nzoia counties, who shared their views as presented in text box 3.

**Table 4:** Motivations for the choice of a particular Pharmacy.

| Reason | Frequency | Percent |
| --- | --- | --- |
| Convenient/close to my home | 13 | 15 |
| Private/confidential care | 22 | 25 |
| Service is good quality | 26 | 30 |
| Service is affordable | 2 | 2 |
| Service is quick/efficient | 6 | 7 |
| The provider is known to the respondent | 10 | 11 |
| Have no alternative | 2 | 2 |
| Others (Specify) | 2 | 2 |

##### Text Box 3

**Views of Pharmacists and Ipas Staff on how MASU project has increased access to MA Services**

###### R (KII-Kisumu)

“yes, I think IPAS has been engaging medical practitioners, pharmacists and making the services accessible to the girls and women through their training and the support we’ve got in terms of the supplies. I would say its helping in reaching out to more girls needing these services.”

###### R (KII-Busia)

“…If I look at what I used to do before in terms of case load, I get to serve many girls from different places that wouldn’t have come before.”

###### R (KII-Busia)

“Yes, many women and girls have been able to access MA since the project started. Generally, I can say there is an improvement in the number of women and girls coming to seek the services of MA.”

###### R (KII- Trans Nzoia)

“It’s widespread because even from the clients we have interacted with, they come from different regions, so when they go back there, they tell the others of the services we offer. When we offer them services, they go there and deliver the information to someone else seeking the same services.”

###### R (KII- Ipas Staff)

Access to MA has improved in the 5 counties following IPAS intervention, but not well expanded yet in the rural areas.

The findings are in agreement with the views of the majority of the participants in the focus group discussions, other users, and youth champions interviewed who identified pharmacies as the place where the majority of girls and women go for safe medical abortion services and that the IPAS MASU project had enhanced access of MA self-use services.

#### 3.3.3 Motivations for the choice of a particular Pharmacy

Reasons as to why the women and girls chose a particular pharmacy and or clinic for MA were asked. The findings are presented in table 4.

The majority of the respondents, 26 (29.5%), reported quality of services as their number one motivation for choosing a particular pharmacy to have an abortion. This was followed by whether a pharmacy can offer private/confidential care, 22 (25%), convenience and or proximity considerations, 13 (14.8%), and whether the provider is known by the user, 10 (11.4%). The same sentiments were shared by the participants in the focus group discussions and users who were subjected to in-depth interviews. This is well illustrated in text box 4.

##### Text Box 4

**Views of FGD and IDI Participants on the motivations behind the choice of a particular pharmacy for the MA services:**

###### R (IDI- Trans Nzoia)

Privacy and confidentiality, and quality services. “I will go back to the same facility in future if I need some other services.”

###### R (IDI- Trans Nzoia)

The pharmacist ensured my privacy and confidentiality. “He respected me and treated me with a lot of dignity. He in fact gave me his contact to call him when I felt or needed something.”

###### (FGD- Trans Nzoia)

Chorus answer: It was very private

###### R (FGD- Kisumu)

Privacy. “I wanted total privacy. So the pharmacist will listen to you make you feel free and comfortable to share your views and issue.”

###### R (FGD-Kisumu)

Pharmacies are okay, because he books you in a private room where you are lone.

###### R (FGD-Kisumu)

You feel safe because you are there only the two of you. You really feel your privacy is guaranteed.

###### R (FGD- Vihiga)

Privacy and how I have been handled. “The provider welcomed me well and made me comfortable and free. And explained to me how to use the drugs and what to do when a complication arose really showed he was skilled enough.”

###### R (IDI-Busia)

I felt my privacy was guaranteed. However, the personnel were very harsh to me.

#### 3.3.4 Packaging of the Abortion Drugs

The majority of the respondents (71.6%) reported that the abortion drugs were sold in a blister pack with all the pills together. Although in many cases, Pharmacists would assist women and girls in correctly administering the abortion drugs, women and girls did not carry home the MA packaging. This was confirmed by participants in the FGDs, and IDIs who could not remember the name of the pills taken because they did not carry the packaging home. This was followed by those who mentioned their abortion drugs or pills were in separate packages – meaning the MA drugs were sold outside the blister pack with both misoprostol and mifepristone separated. The finding was corroborated by some users interviewed who confirmed that the tablets were given to them separately. Nearly 9% reported they were given individual pills without packaging. This information is presented in figure 1.

**Figure 1:**
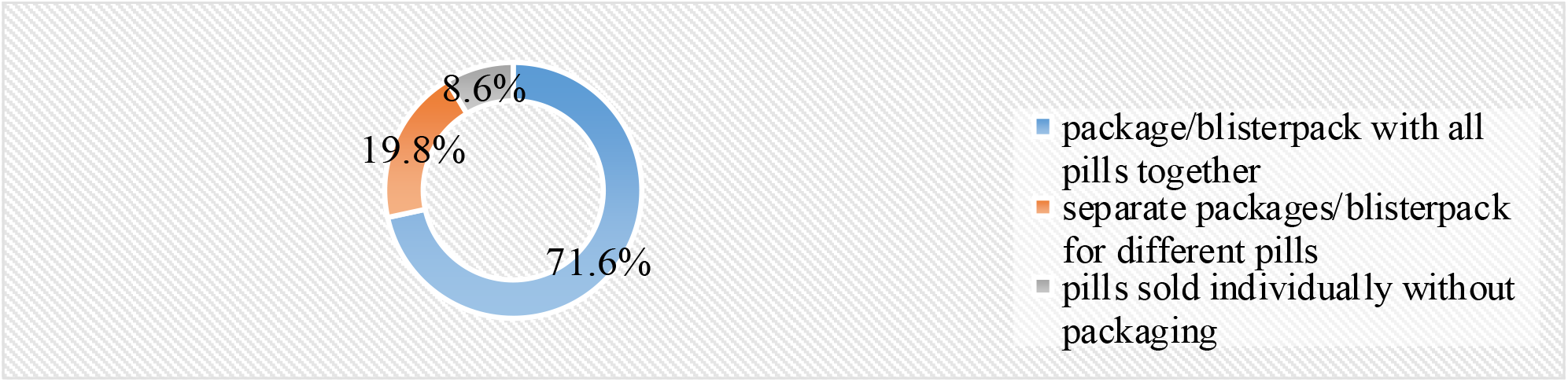
Packaging of Abortion Drugs.

#### 3.3.5 Awareness of the type of Pill taken

When probed if they are aware of the type of pill they were given, 60% of the respondents answered in the affirmative. Similarly, in the focus group discussions, and in-depth interviews of users it was observed that the majority of the users knew the specific pill used.

The findings on the awareness among the users on the type of pills used were supported by the views of the various users interviewed across the five counties as presented in text box 5.

##### Text Box 5

**Views of various IDI Respondents on the Type of Pill Taken**.

- ***R (IDI-Busia):*** R: pills, I don’t know the name actually but. I was given some pills and then you swallow.
- ***R (IDI-Busia):*** starting with the name of the pill, to be honest I can’t remember, I didn’t know it until now I don’t know. My aim and target were just one, to get the drugs take them, it gets flushed out then am done.
- ***R (IDI- Vihiga):*** R: yes, combi pack

#### 3.3.6 Administration of Medical Abortion Pills

##### a) Mode of Taking Abortion Pills

Respondents were asked to report how they took the pills. The majority of the respondents, 53 (60%) reported that they took the pills both together and at separate times. Those who reported at separate times were 25 (28%). Figure 2 presents these findings which were in line with the views of various participants in the focus group discussions and IDI respondents across the counties of study, some confirmed that they were asked to take the pills together, as others took them at separate times.

**Figure 2:**
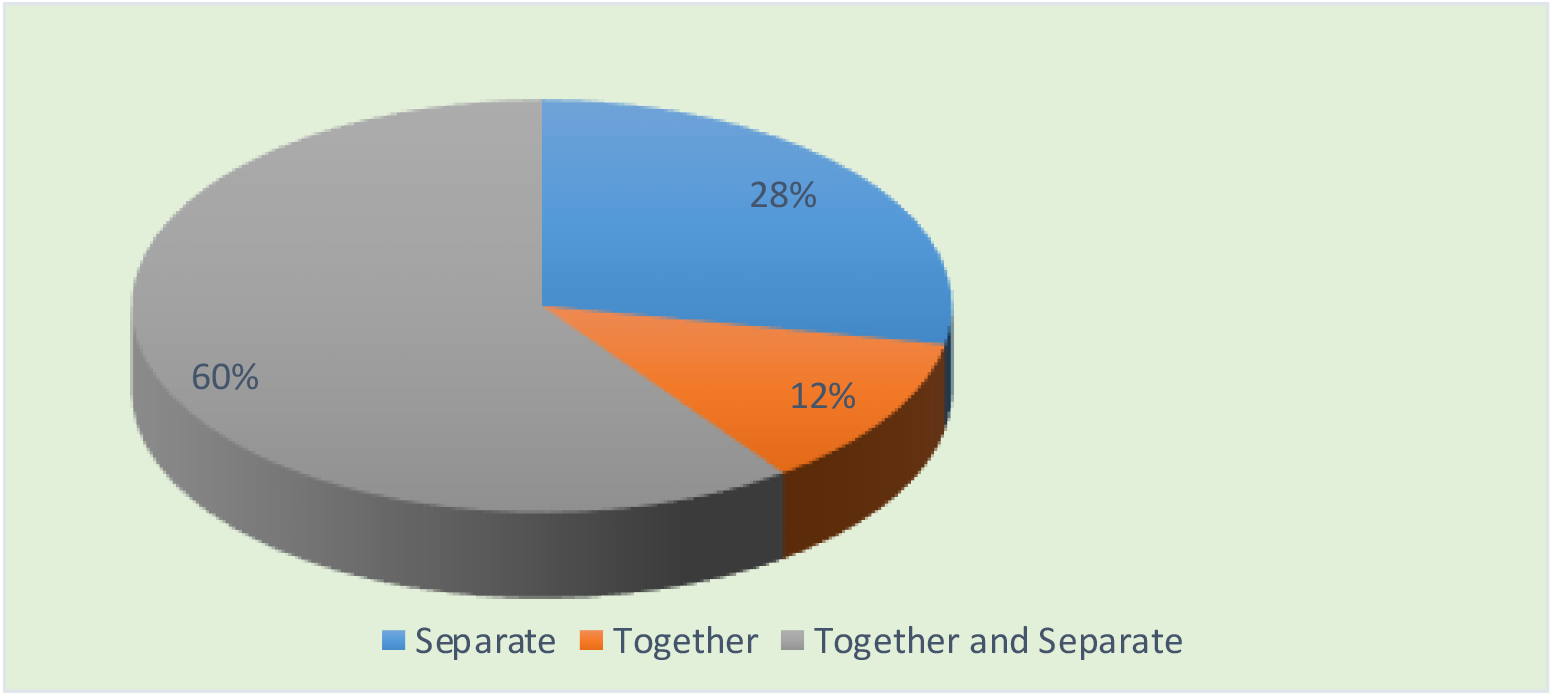
Mode of taking Pills.

##### b) Periods for taking the Pills

When it comes to the number of hours women and girls seeking MA were to take the abortion pills, the majority (44) indicated 24 hours (within a day), which is the recommended interval for the *combi pack*, which is widely used and known for a higher level of efficacy. This was followed by those who reported 12 hours (half-a-day) also within the recommended interval, and 48 hours (2 days) at 7 and 6 individuals respectively. On the same note, various participants in the FGDs and IDI respondents also confirmed there was a lack of uniformity on the time it took them to complete the dosage of the pills with some indicating 24 hours (recommended), others 48 hours (still okay but in case of any delay may be ineffective), and some half a day, 12 hours. Figure 3 shows these results The views of various users on the period for taking the pills are presented in Text Box 6.

**Figure 3:**
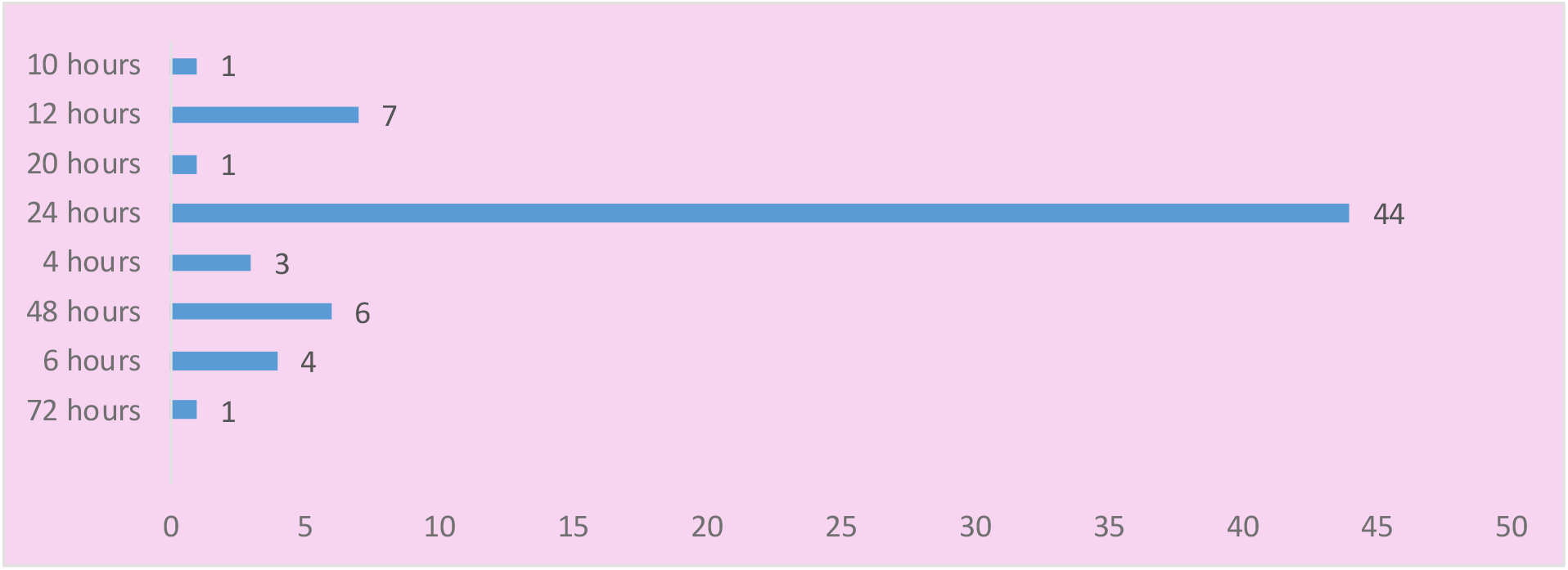
Periods for taking the Pills

###### Text Box 6

**Views of various IDI Respondents on Periods for taking the Pills**

###### R (IDI-Busia)

for the services I was satisfied with them, what I didn’t have interest to know was the medication because my worry was to get helped for it to come out. The tablets he gave me were 5, I went took one with water that was on a Thursday then on Saturday he told me to place the others under the tongue.

###### R (IDI-Kisumu)

I went there and as told I got prepared psychologically then I was given the one tablet and told to come after two days. Was told to take lots of water within the two days. After the two days come back and given the four tablets that I placed under the tongue. That was it.

###### R (IDI- Vihiga)

the experience is that I didn’t expect that’s how to go about the medication. You know those drugs are that some you put under the tongue another you insert down in the vagina another you are told to swallow then after 24 hours you start inserting the one for below. I just thought it’s a one-day issue then the following day you are okay, unlike knowing it has those two days

##### c) Route of Administration of the Pills

Respondents mentioned swallowing and placing them below the tongue as the common route of administration (sublingual route) for abortion with pills. The sublingual route of administration had the highest utilization especially by clients using combi pack at 41 (49%) and 13 (16%) for misoprostol respectively. Other respondents mixed both sublingual and vaginal routes in the process of administration. Figure 4 highlights the findings.

**Figure 4:**
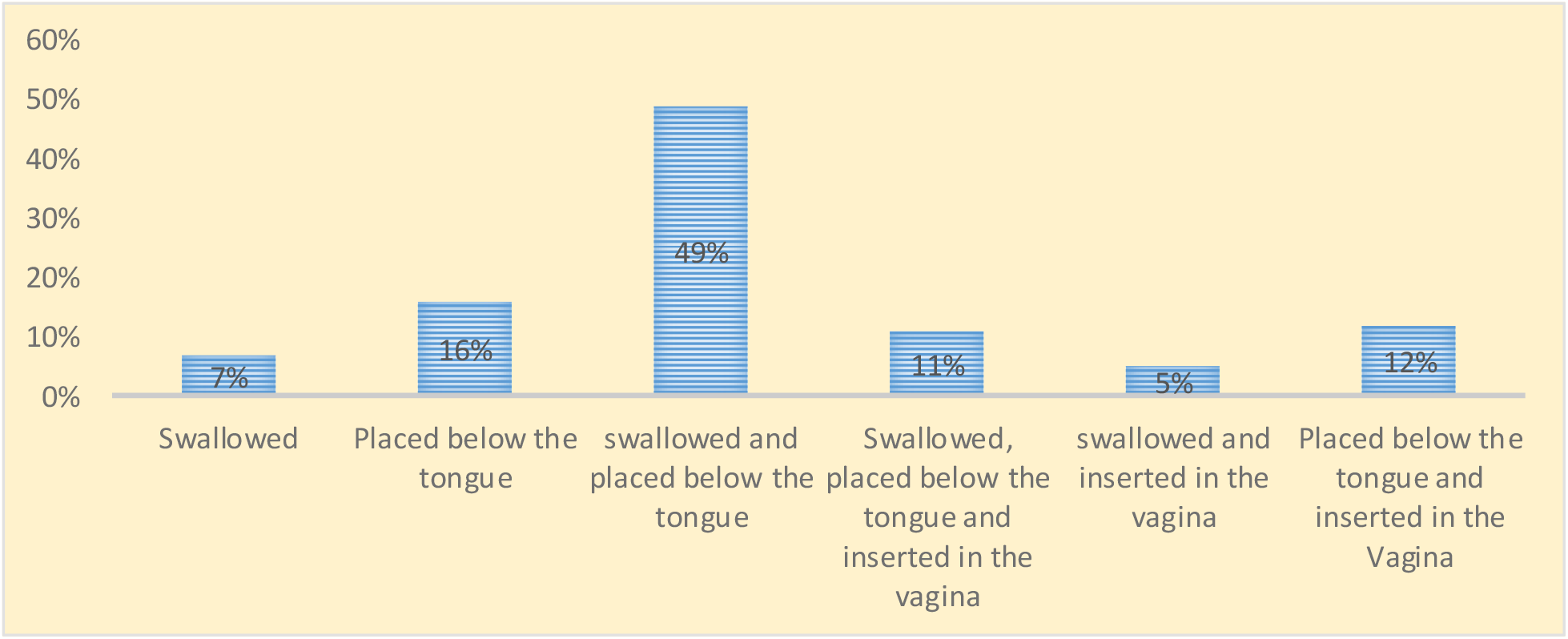
Administration of Abortion Drugs.

The findings corroborate what emerged from the Focus Group Discussions where some participants indicated to have taken the first tablet at the pharmacy, then went back the following day to take the remaining four to avoid being seen with the tablets, either at the home or in the hostels. Others reported having swallowed one tablet, then either placed the remaining four below the tongue or inserted them in the vagina as instructed by the pharmacist. These views were the same as those expressed by the various participants in the IDIs interviews.

#### 3.3.7 Awareness of the gestation period

A total of 69 (78%) users of medical abortion reported being aware of the gestation period within which they can safely use medical abortion pills. However, some pharmacists interviewed observed that some users are honestly not aware of the gestation period, whereas others lie about their gestation period if it is past the window period and they require the services. Other users deliberately give lower periods hoping it would reduce the cost of service. The views of respondents on awareness of the gestation period are presented in Text Box 7.

##### Text Box 7

**Views of various FGD, KII, and IDI participants, and Pharmacists on user’s awareness of the Gestation Period**.

###### R (IDI-Busia)

three weeks and below, this is the time you it’s safe for a woman to procure and it won’t affect them, but beyond three weeks there could be some dangers.

###### R (IDI-Siaya)

R: it should be 10 weeks or 70 days after your last menstrual period

R: yes, she can over bleed, cramping, she can die incase not all products are removed

###### R (IDI – Trans Nzoia)

R: It should be around two months

###### R (FGD -Kisumu)

I think it’s that moment that you are pregnant, but it has swollen yet, it’s still low in the belly is the right time to terminate, like it’s not beyond two months that’s safe to terminate.

###### R (KII-Siaya, Pharmacist)

There are still some challenges, basically I can’t really pinpoint a major challenge only that there are women who come with a bigger gestational age that we must refer them. They also insist that since I am the first person, they have come to them insist I have solved the problem for them.

###### R (KII-Ipas Staff)

Inability to accurately estimate the gestation period, some users lie about the gestation hoping to lower cost of medication

We further analyzed the measures of central tendency for the number of weeks when it is considered safe to use MA. The median number of weeks for safe MA abortions stood at 5 weeks.

#### 3.3.8 Complications when Having an abortion

Of the respondents (users of the medical abortion services) 65 (74%) reported no medical complications after an abortion procedure. Participants in the FGD, pharmacists and youth champions also reported cases of complications during the administration of safe medical abortion self-care. The most common form of complications identified by respondents were excessive bleeding, swollen eyes, severe headache, and, extreme cramping.

#### 3.3.9 Management of Abortion-Related Complications

Respondents who had developed abortion-related complications reported being given painkillers and antibiotics as the most sought recourse when they developed post-abortion complications, 17% reported of those with complications visited a clinic for professional medical services to handle such complications. This is highlighted in figure 5.

**Figure 5:**
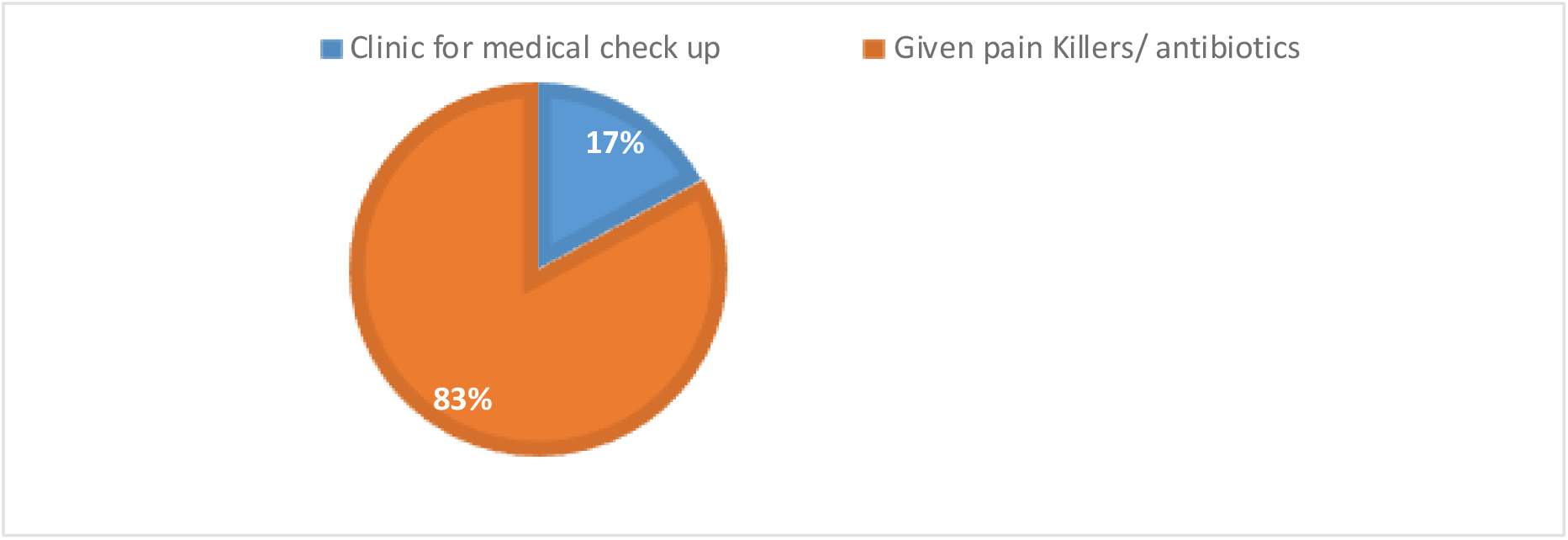
Management of post-abortion-related complications.

##### Text Box 8

**Views of various FGD participants on how they managed complications in the post-abortion period**.

- ***R (FGD-Busia):*** “I would say it’s okay, but one should know they can even die. Like for me, I could have died from the procedure. Being inserted the plastics is not an easy way. It was only that I didn’t have another way, but it also helps.”
- ***R (FGD-Kisumu):*** “For me he told me, since he had given me the drugs and procedures to use, I should be free to call him and inquire in case any complications arose. I should fear to call and talk to him.”

The above findings have been corroborated by the FGDs as presented in text box 8.

#### 3.3.10 Traveling Time to the Pharmacy/clinic offering Abortion Services

The majority of the respondents 61 (74%) reported traveling for less than 30 minutes to access facilities offering medical abortion services. Those who reported traveling for between half an hour and two hours were 16 (19%). One respondent reported traveling for a day to receive abortion services. This is presented in figure 6. The findings were in agreement with the sentiments expressed by the various FGDs and IDIs participants who also confirmed traveling for about 30 minutes to a maximum of 2 hours to access facilities that offer safe MA self-use services.

**Figure 6:**
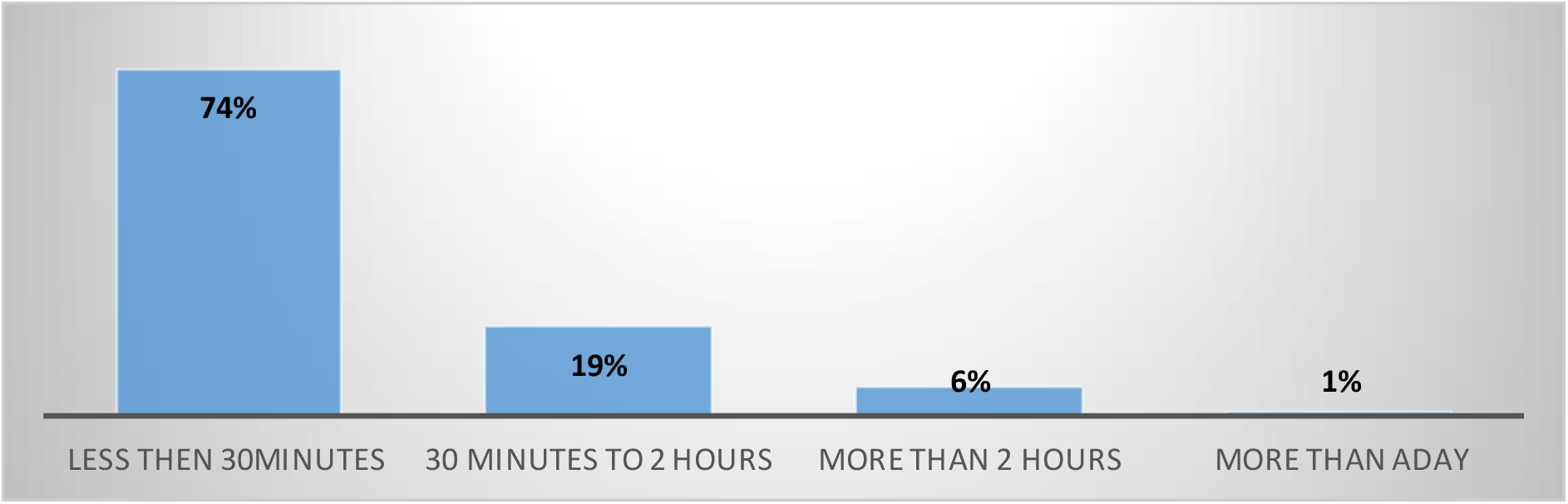
Time taken to Travel for abortion services.

The respondents were further probed on how long it took them to get medical abortion services (waiting time). Those who said they waited for 30 minutes (half an hour) were 68 (82%). This has been followed by those who reported that they waited for between half an hour and two 2 hours at 11 (13%). Only 5% indicated that they waited for more than 2 hours to have a medical abortion. Figure 7 depicts these findings. The same findings were corroborated with the IDIs and FGDs where the majority confirmed waiting time for safe MA services at the pharmacies to range between 30 minutes to 2 hours on average.

**Figure 7:**
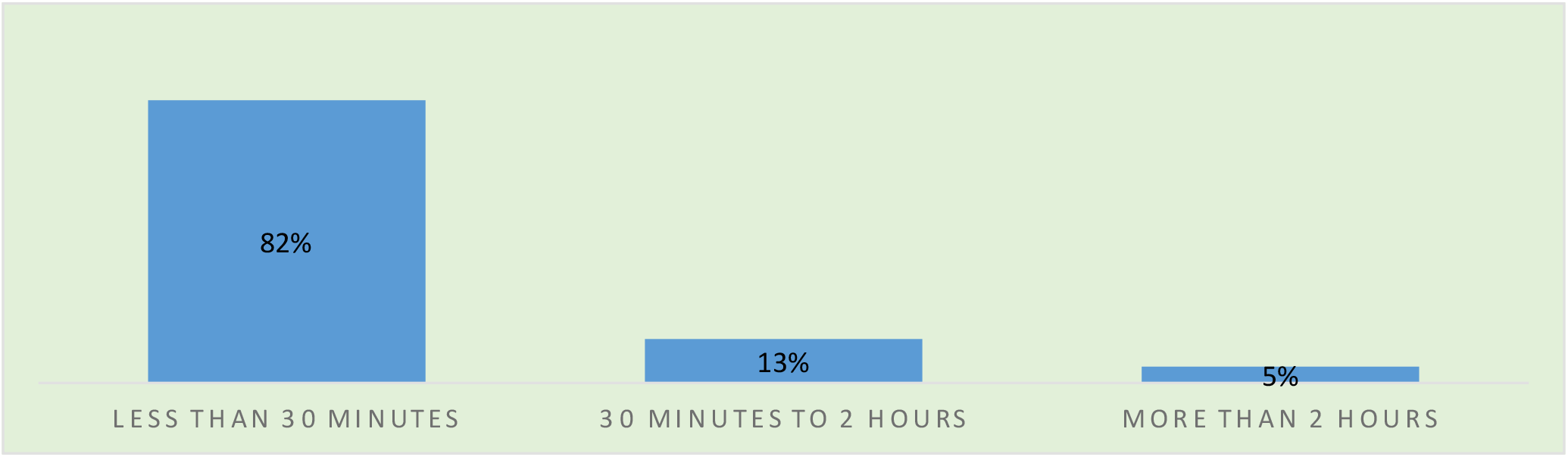
Waiting time at the Pharmacy before MA Services.

#### 3.3.11 Pain Management Drugs

Users of medical abortion were also probed whether they were given pain management drugs when they sort for the abortion procedure. 78 (89%) reported having been given pain-relieving medications. On the other hand, 10 (11%) reported not having been given the medication. The views of pharmacists, FGDs, and IDI respondents are expressed in text box 9.

##### Text Box 9

**Views of various FGD participants, and IDIs on Pain Management Drugs**.

- ***R (IDI-Trans Nzoia):*** after she had inserted the other 4 now, this was 24 hours after using the one of the tongues. Like, she put one under the tongue, waited for 24 hours, took a painkiller then later inserted the 4 in the vagina.
- ***R (FGD-Busia):*** I was given painkiller bruffen, but on that Saturday when I used the ones for the vagina on Sunday morning, I woke up my eyes were all of them swollen, I called him. He told me to relax it will be okay. After two three hours they went back to normal
- ***R (FGD-Busia):*** I wasn’t given any painkiller, I was given those drugs and was told I must follow instructions and it will be fine, yes it was all fine though felt some little stomach cramps, but I didn’t go back coz I didn’t want to be seen there again
- ***R (FGD-Kisumu):*** For me, I can say the pain was too much, but he offered me painkillers. He said I take them after six hours. You know you have to follow instructions given by the doctor, so for the six-hour period I was really in pain.

Those who indicated they were not given any pain management drugs in the post-abortion period were asked to state the reasons why they believe this is happening. A majority (50%) of the respondents reported that they don’t know. Those who said they did not ask for any pain management drug were 20%, same as those who indicated they received a piece of advice not to take any medication apart from what was prescribed by the pharmacy personnel. These findings are shown in figure 8.

**Figure 8:**
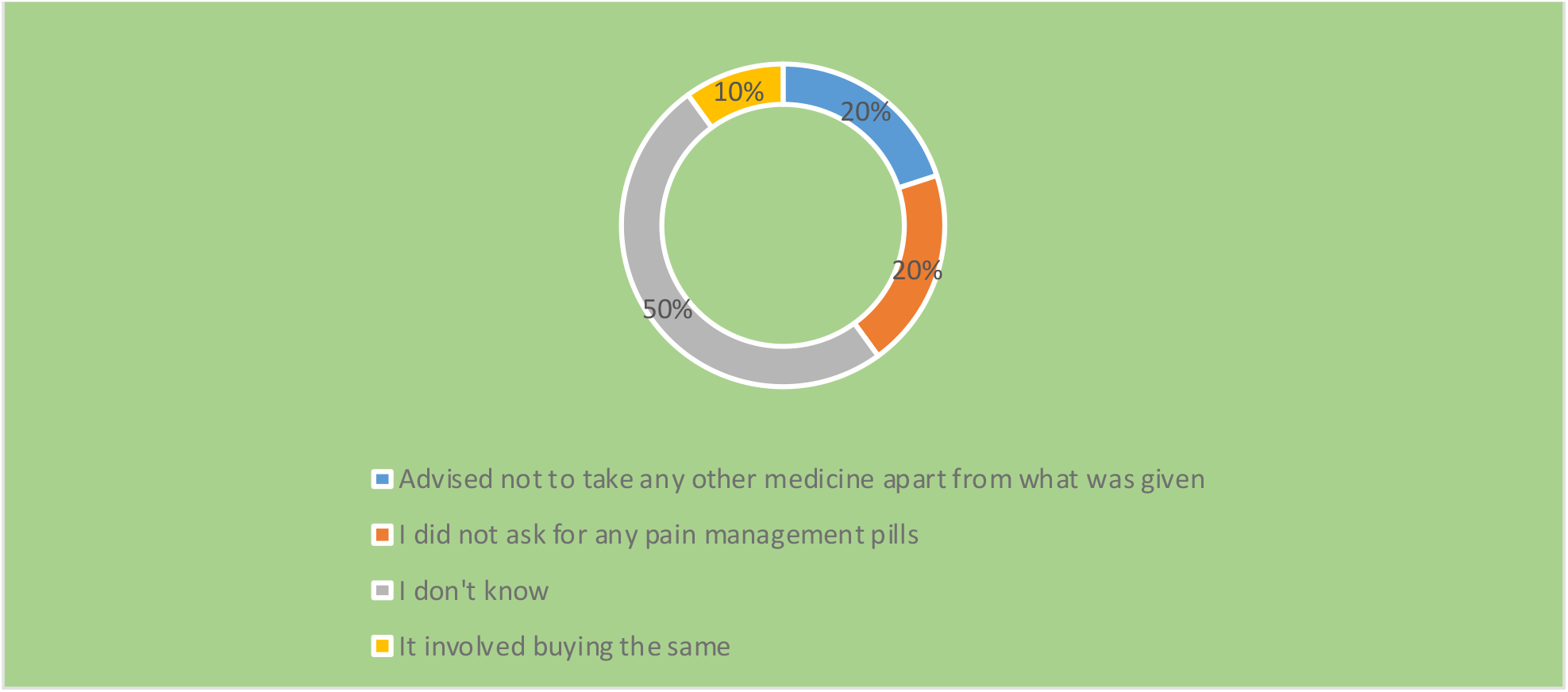
Why Pain Management Drugs were not given.

#### 3.3.12 Days the pharmacist took to make a follow-up on the MA users

The number of days it took the pharmacists to follow up on the women and girls who had sought a medical abortion from their facilities was also probed. The majority of the respondents (56%) indicated it took the pharmacist who served them between zero to three days (which is the recommended period to address any complication that may arise) to contact them wanting to know how they are doing and if there is any problem requiring their attention. Those who reported a follow-up period of between four to seven days and no follow-ups were 27% and 10% respectively. Figure 9 illustrates these findings.

**Figure 9:**
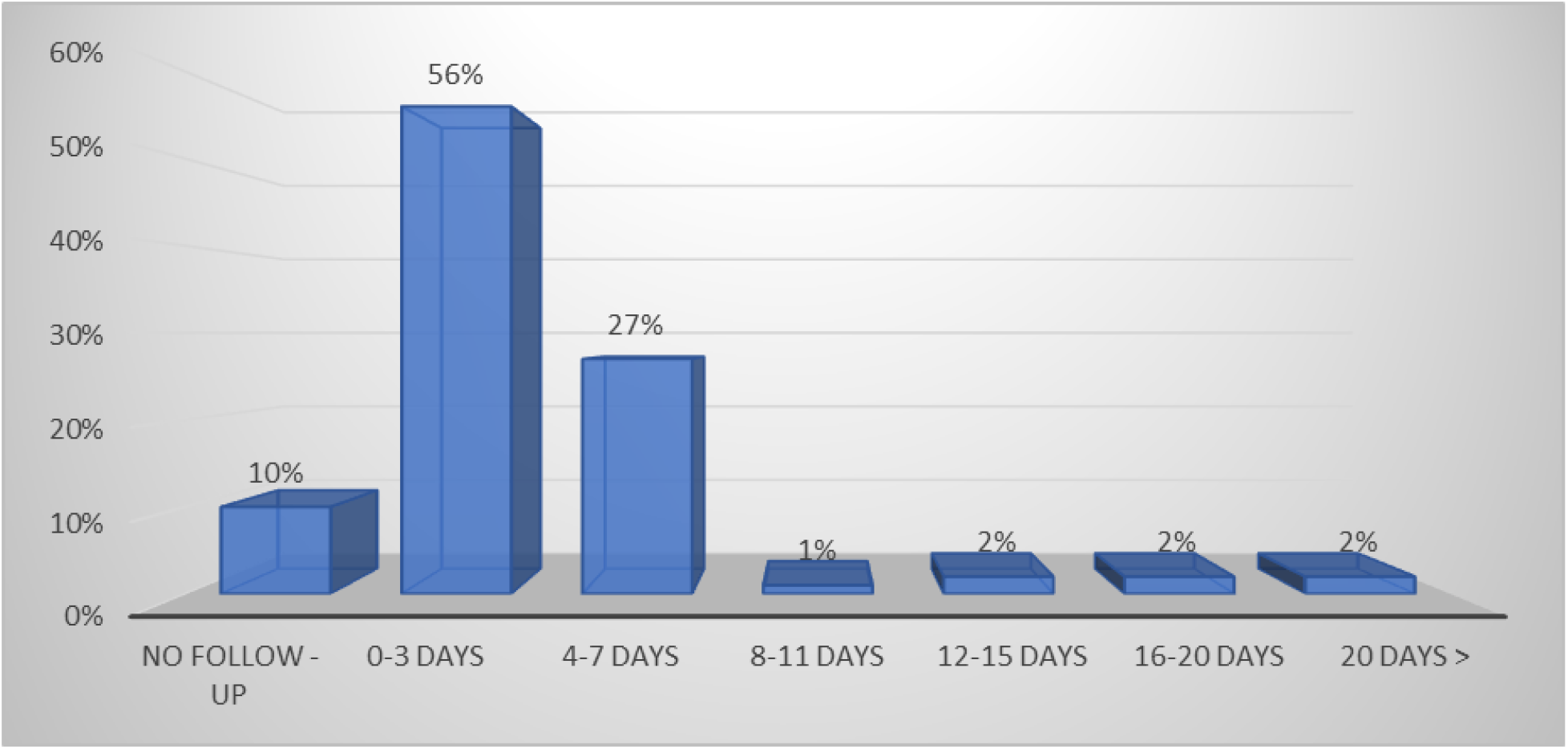
Number of Days the pharmacies took to make follow-ups.

### 3.4 Contraception After Abortion

The majority of the respondents (90%) reported that they were asked whether they want to delay and prevent any future pregnancy given the abortion experience. The same proportion of respondents, 90%, also reported being given information on the available contraception after the abortion procedure.

The views of the various users interviewed on whether they were given information on the available contraceptives in the post-abortion period are presented in Text Box 10.

#### Text Box 10

**Views of various IDIs and KIIs whether the Information on Contraceptives Available was Given**

##### R (IDI-Trans Nzoia)

Yes, that there is the three months injection, pills, the one they put in the arm Since my daughter is still in school, I decided her be put implant, the three years one.

##### R (IDI-Trans Siaya)

yes, it was comprehensive. He advised me very well, he told me the injection can affect one, that it can make one fail to conceive, and the implant if one can feel the side effects can go back and have it removed. The information was good.

##### R(KII-Busia)

I take them through the counselling on contraception. I tell them of the options that we have and how they work out, the side effects, how they work out and what to expect. I always give them two sets of counselling which I feel I do my best.

#### 3.4.1 Opportunity to ask questions concerning different contraceptives available in the postabortion period

Women and girls accessing medical abortion were also asked whether there was room to ask questions to the personnel at the pharmacy and or clinics concerning different contraceptive methods during service delivery. 88% stated that there was room to pose such questions.

#### 3.4.2 Given Contraceptives during the post-abortion Period

Nearly half (47%) of the users of medical abortion reported they were not given post-abortion contraceptives. On the other hand, 53% reported having been given contraceptives. The findings are presented in figure 10.

**Figure 10:**
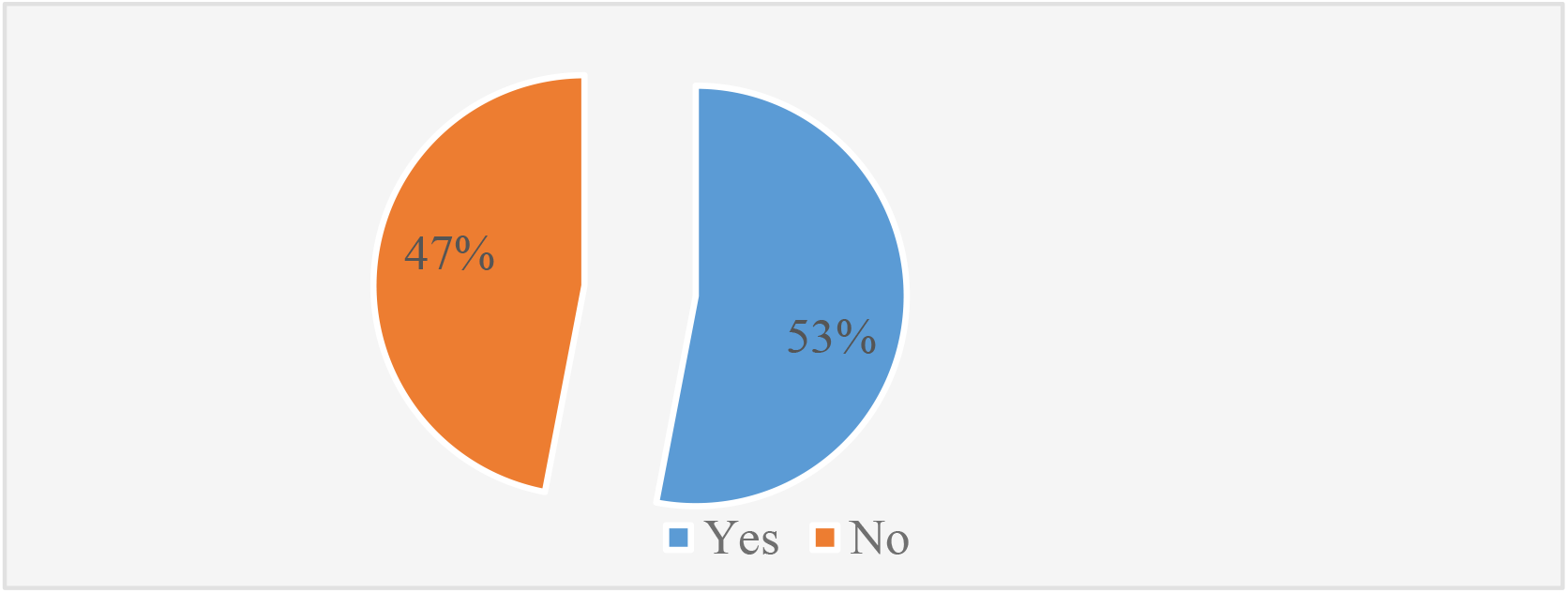
Given Post-Abortion Contraceptives.

The views of various IDI and KII respondents on whether they were given Contraceptives during the post-abortion Period are presented in Text Box 11.

#### 3.4.3 Contraceptives MA Users were Counselled on by the Pharmacists

##### Text Box 11

**Views of various IDIs and KIIs on whether they were Given Contraceptives during the post abortion Period**

###### R (IDI-Busia)

I was advised to take the injection, but I was not ready for that. I took time then after that I went back to pick it.

###### R (KII-Busia)

Now the problem with these girls is that after giving them all this information, they tell you let me go and come back, but they never return. There is still that fear of using contraception, especially for the school going girls

###### R (KII-Siaya)

Yeah, that I was now told it’s an option, because there are some who feel eeh I haven’t given birth and these FPs parents sometimes say it can destroy something.

As regards the contraceptives the MA users were counseled on when they sought a medical abortion in the pharmacies, the majority of the respondents 16 (21%) reported they were counseled on the use of pills. This was followed by injectables, 14 (19%), and implants, 10 (13%). On the other hand, 15 (20%) of the women and girls reported being counseled on pills, emergency contraceptives, injectables, IUCD, and implants but not on female sterilization. This is presented in figure 11.

**Figure 11:**
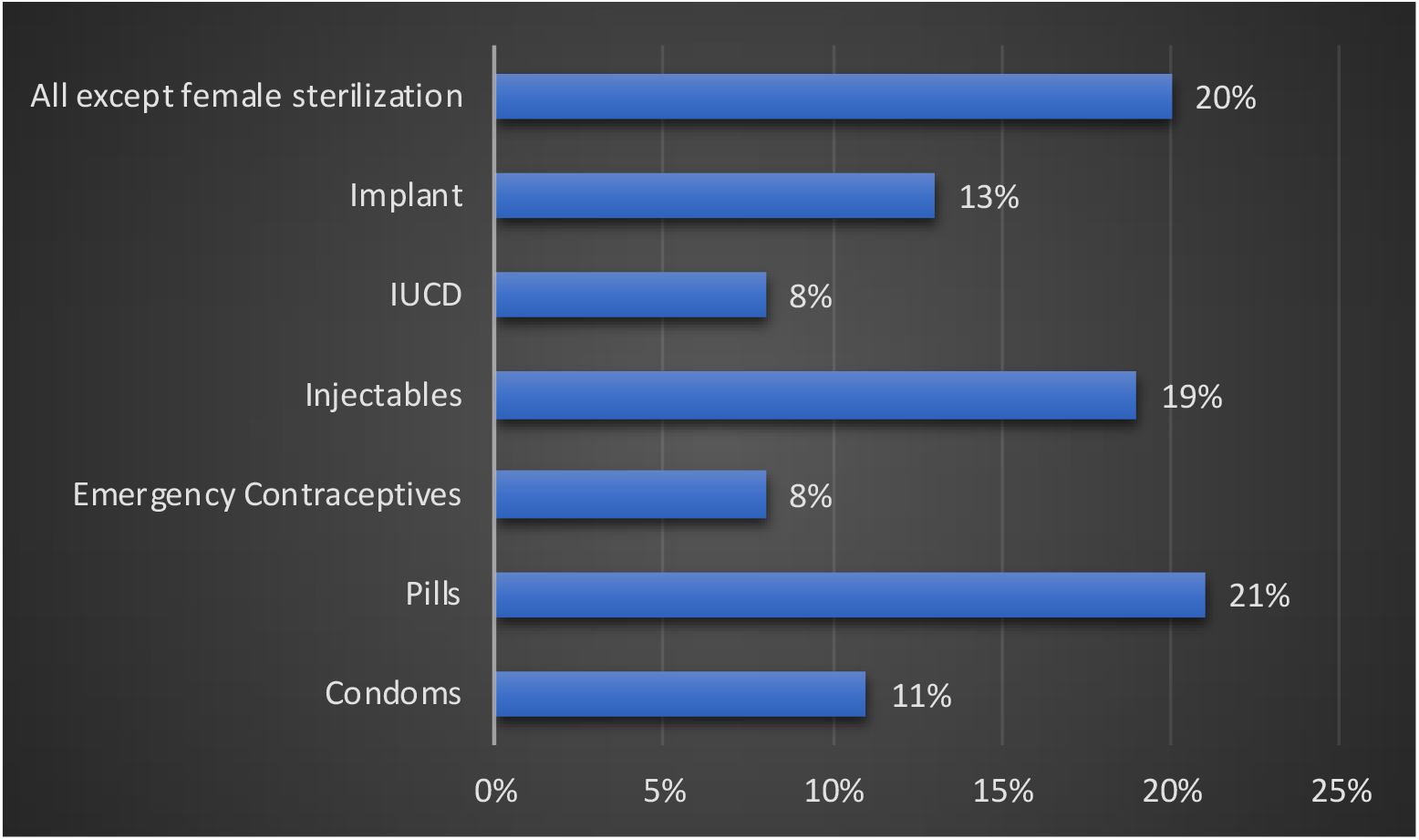
Contraceptives MA Users were Counselled on by the Pharmacists.

#### 3.4.4 Type of Post-abortion Contraceptives given

On the type of contraceptives given in the post-abortion aftermath, the majority of the respondents 24 (27%) reported having been given injectables. This was followed by pills, implants, and Condoms at 8 (9%), 6 (7%), and 5 (6%) people respectively. IUCD had only one respondent. It is important to determine the effectiveness of each and the associated side effects and or the risks that each exposes the users to and their resolve to have children in the future. These findings are illustrated in Table 5.

**Table 5:**
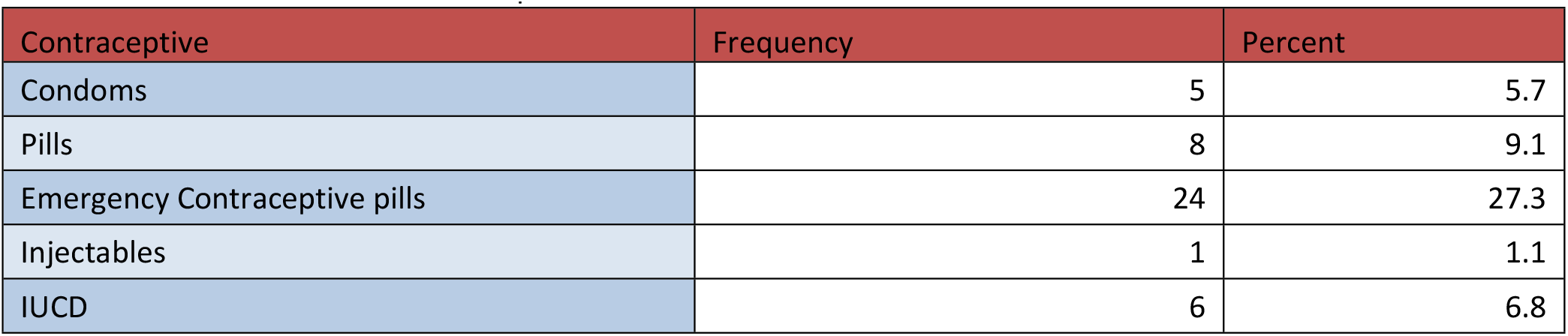
Given Post-Abortion Contraceptives

| Contraceptive | Frequency | Percent |
| --- | --- | --- |
| Condoms | 5 | 5.7 |
| Pills | 8 | 9.1 |
| Emergency Contraceptive pills | 24 | 27.3 |
| Injectables | 1 | 1.1 |
| IUCD | 6 | 6.8 |

The FGDs also came up with the same findings as shown in Text Box 12:

#### 3.4.5 Contraceptive (s) received wanted

Women and girls who were given post-abortion contraceptives were asked to state whether the contraception received is what they wanted. A majority (53%) reported that yes, it is what they wanted.

##### Text Box 12

**Views of various FGD participants on the type of post-abortion contraceptive given**.

###### R (FGD-Busia)

- “I took the three months injection twice now.”
- “I was given that one of five years.”
- “for me she wanted to give me the injection immediately and I refused.” … “Wanted to finish the procedure first.”

###### R (FGD- Trans Nzoia)

“The pharmacists offered contraceptive counselling.”

#### 3.4.6 Why the preferred Contraceptive wasn’t given

More than half of the respondents (55%) indicated lack of the contraceptive preferred at the pharmacy attended was the primary reason why they didn’t get the contraceptive they wanted. This is followed by being pressured to accept the available contraceptive by the pharmacy personnel at 18%. Additionally, 14% of the respondents reported that they have no idea about the type of contraceptive they should use/want. Further, 11% of the respondents mentioned that the contraceptive offered wasn’t their preference. These findings are shown in figure 12.

**Figure 12:**
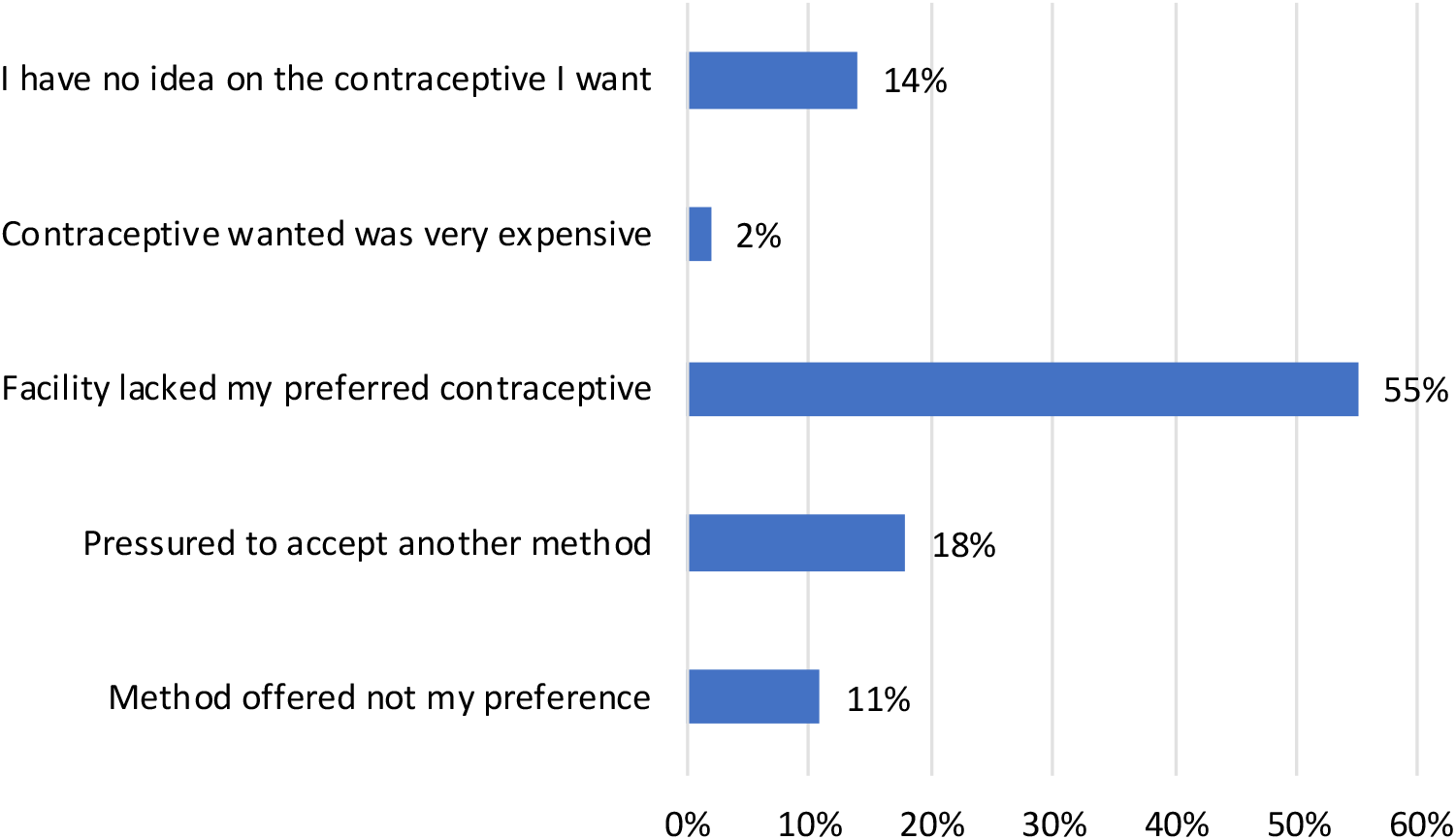
Why preferred Contraceptives were not given.

#### 3.4.7 Pressured to accept a particular Contraceptive method

The Majority (86%) of the users of medical abortion reported being pressured to accept and use a particular type of contraceptive against their wishes. However, it remains unclear on what premise such pressure was based on the point of view of the medical abortion provider. Many providers suggested methods to users to increase their sales and did not provide adequate information to guide the users in selecting viable methods.

Text Box 13 presents the views of various FGD and IDI respondents on whether they were forced to take a particular contraceptive at the pharmacy.

##### Text Box 13

**Views of various FGD, IDIs, and KIIs on whether they were forced to take a particular contraceptive**

###### R (IDI-Busia)

I was advised to take the injection, but I was not ready for that. I took time then after that I went back to pick it. He told me to choose for myself

###### R (IDI-Trans Nzoia)

I opted for the three months injection. I didn’t feel pressured to choose the injection and it was available for use when I needed it.

#### 3.4.8 Advise on Possible Side Effects of Contraceptives

The majority of the respondents reported that they were well informed on the side effects of the contraceptive method given. This information was obtained from experiences with peers and friends. Figure 13 depicts the findings.

**Figure 13:**
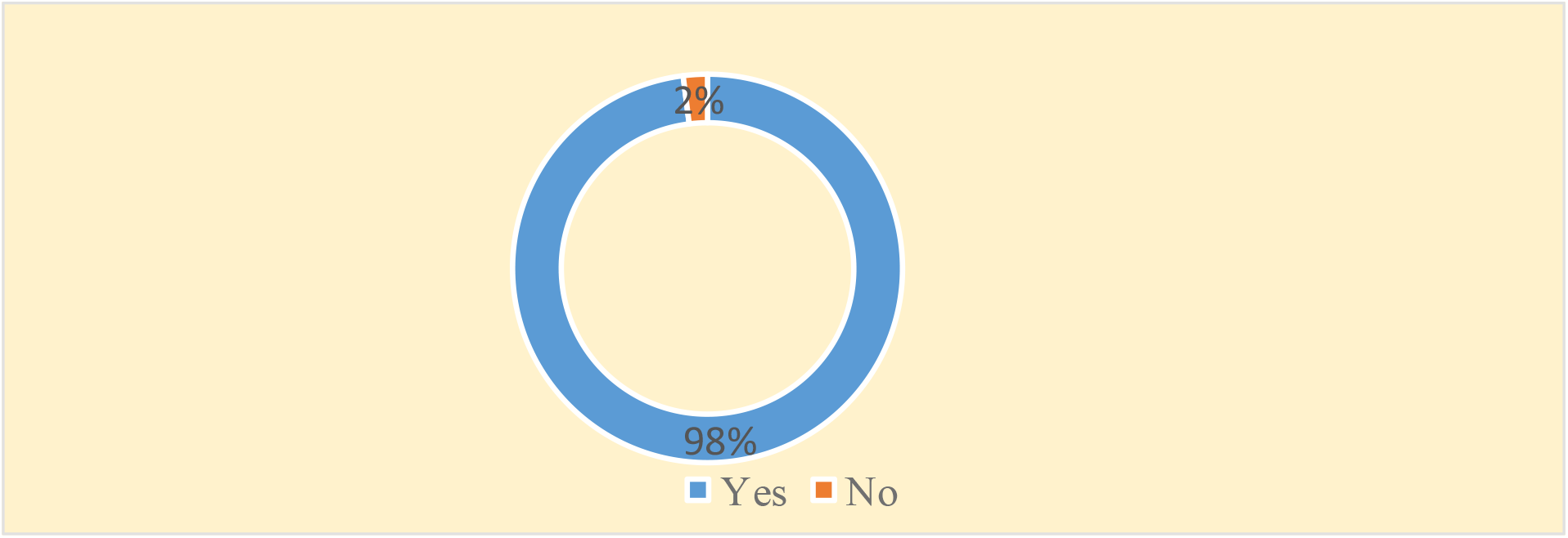
Advice on Possible Side Effects of Contraceptives.

### 3.5 Medical Abortion Client Feedback and Satisfaction

The table below summarizes women’s and girls’ feedback on some issues touching on their perceptions of the Pharmacists they sought medical abortions from. Nearly 99% reported they were given confidential care. What should be of keen interest is the 22% of the respondents who reported that the pharmacist’s personnel who handled them did not support their decision to terminate their pregnancy. These are presented in table 6.

**Table 6:** Summary of MA user’s Perceptions of the services offered by the Pharmacists.

| Variable | Frequency (n) | Percent (%) |
| --- | --- | --- |
| **Trust to be given confidential Care** |  |  |
| Yes | 82 | 98.8 |
| No | 1 | 1.2 |
| **Would you return to the facility** |  |  |
| Yes | 79 | 95.2 |
| No | 4 | 4.8 |
| **Treated with respect and dignity** |  |  |
| Yes | 82 | 98.8 |
| No | 1 | 1.2 |
| **Recommend facility to a friend** |  |  |
| Yes | 80 | 96.4 |
| No | 3 | 3.6 |
| **Supported in Pregnancy termination** |  |  |
| Yes | 66 | 79 |
| No | 17 | 21 |
| **Needs and concerns listened to during a visit** |  |  |
| Yes | 82 | 98.8 |
| No | 1 | 1.2 |

The pharmacists also self-appraise themselves in terms of the adequacy of the services they offer to women and girls. They majorly reported that the services offered are adequately reflected by the positive feedback they normally get. The responses have been presented in Text Box 14 verbatim.

### 3.6 Average costs of Medical Abortion

#### Text Box 14

**Views of various Pharmacy participants on the adequacy of the services offered within their facilities**

##### R (IDI-Busia)

For those who have come to me none has left without the support they needed. Also, when I do the follow-up, for those who experience any complications I’ve been able to successfully refer them for more help and they always bring positive feedback. I feel I have done what am supposed to do adequately.

##### R (IDI- Trans Nzoia)

“Yes, the services are adequate due to the feedback that I normally get. You serve a client she goes aways and comes back it tells you the services were good; they didn’t have any challenges. With you feel it was adequate.” R (IDI-Siaya):

▪ “Yes, I feel they are adequate being that the number of clients who come back with failed abortions are none.”
▪ “Recently I met a lady with septic abortion…So a week later the lady was brought here at night with septic abortion. I did to her an MVA and put her on medication, IV drugs. That means there’s a wide gap out there as people still come reporting of septic abortion.”
▪ “Many people should be trained, and many pharmacies reached to dispense MA drugs better.”

##### R (IDI- Kisumu)

Since we started with IPAS…the quality has gone high and its been improved greatly. This is because, we were taken through a protocol which is quite important and what happens after following the protocol, the follow-up bit, you find that we found the missing link that used to be missing. There have been improvements

When it comes to the cost incurred for an abortion procedure at the pharmacy/clinics/chemists attended, the majority of these users (30) reported an average cost of Ksh. 2100 to 3000. This was followed by a cost range of 1000 to 2000 and Ksh. 3100 to 4000. The median abortion cost stood at Ksh. 3000. The findings are presented in figure 14.

**Figure 14:**
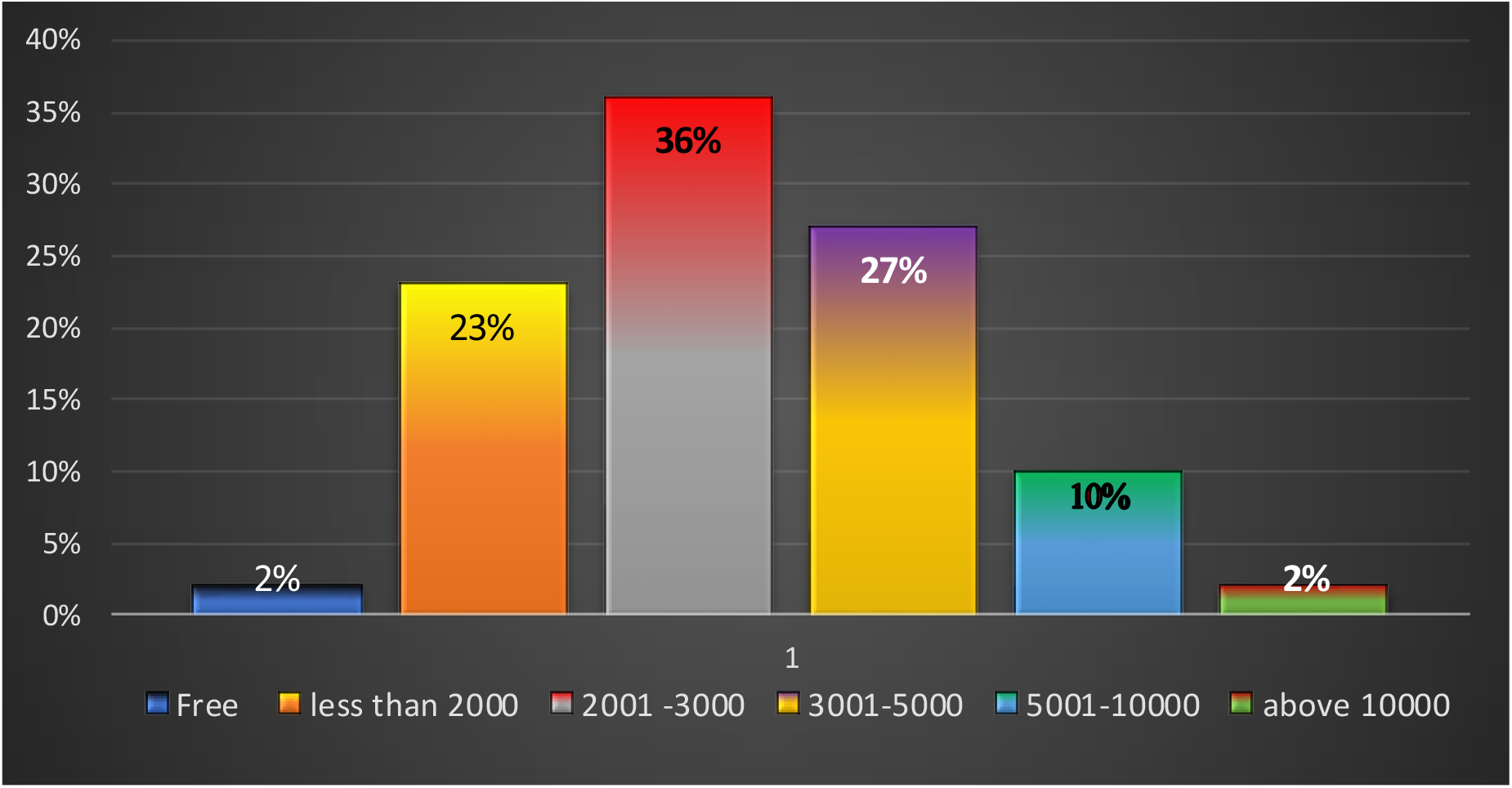
Cost of Medical Abortion.

Text Box 15 presents the views of various IDI and FGD respondents on the cost of safe medical abortion services. We did a further analysis by cross-tabulating abortion costs and County of residence. From the analysis, Vihiga County has the highest number of people who indicated they paid an average of Kenya Shillings between 5,100 and 7,000. At the same time, the County (Vihiga) is the only county with individuals indicating they paid from Ksh. 9,000 and above for medical abortion. This is well illustrated in table 7.

**Table 7:** Cross-tabulation of abortion by Counties.

|  | Abortion Costs |  |  |  |  |  |  |
| --- | --- | --- | --- | --- | --- | --- | --- |
|  | Free | 2000 | 4000 | 6000 | 8000 | 9,500 | 11500 |
| County |  |  |  |  |  |  |  |
| Busia | - | 6 | 6 | - | 1 | - | - |
| Kisumu | 1 | 12 | 2 | 1 | - | - | - |
| Siaya | 1 | 10 | 2 | - | 1 | - | - |
| Trans Nzoia | - | 14 | 3 | 1 | - | - | - |
| Vihiga | - | 10 | 11 | 3 | 1 | 1 | 1 |
| Total | 2 | 52 | 24 | 5 | 3 | 1 | 1 |

#### Text Box 15

**Views of various FGD and IDIs on the Cost of Safe Medical Abortion Services**

##### R (IDI-Trans Nzoiya)

The cost is affordable. I paid ksh. 3000

##### R (FGD-Busia)

R1 I found it to be affordable because I needed help.I was charged Ksh.2500

R2: 3500; R2: 1500; R3: 3000

However, the pharmacists during the In-depth interviews narrated that their costs of operations when dispensing MA services have drastically come down due to the presence of reliable drug suppliers through the MASU project. This is depicted in Text Box 16.

#### Text Box 16

**Views of various Pharmacist participants on the cost of MA because of MASU project**

##### R (IDI-Busia)

- “First is access, IPAS to some extend provide us with the commodities, the pills. This makes it easy to give it out at a fair cost unlike before where we used to get them at a higher cost thus passing it to girls who at times could not afford it and end up going to back streets to access the services hurting themselves using unsafe methods.”
- ***R (IDI- Kisumu):*** “…since we started our partnership with MASU clients have got quality services owing to the constant Continuing Medical Education (CMEs) we have and trainings, and even the support in regard to how we follow-up these girls in terms of data and airtime.”
- ***R (IDI-Siaya):*** “Before I used to buy the drugs at 700 shillings combi-pack but now there’s a chemist in Kisumu selling them at 150 shillings. This means when I access drugs at a cheaper price the end user will also get them a fair price.”

#### 3.6.1 View on Cost of Medical Abortion Self-Care Services

When probed further on their feeling about abortion costs, 38 (43%), people reported the cost as affordable, 22 (22%), reported it as expensive, while 10 (11%), people averred that it was very expensive. This is well depicted in Table 8.

**Table 8:**
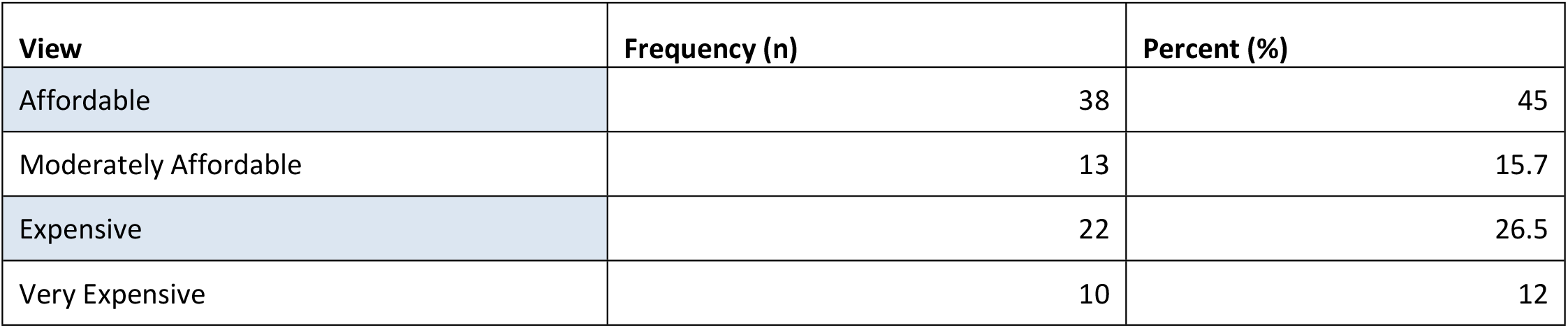
Views on Abortion Costs.

| View | Frequency (n) | Percent (%) |
| --- | --- | --- |
| Affordable | 38 | 45 |
| Moderately Affordable | 13 | 15.7 |
| Expensive | 22 | 26.5 |
| Very Expensive | 10 | 12 |

**Text Box 17 presents the views of various respondents on the cost of safe medical abortion services**.

##### Text Box 17

**Views of various Pharmacist participants on the cost of MA as a result of MASU project**

###### R (IDI-Busia)

R1: I paid ksh. 3000 I found it to be affordable because I needed help. R2: I was charged 2500. Since I needed help, I didn’t feel the weight of the cost

###### R (KII-Ipas Staff)

Ipas MASU project has been able to reduce the cost of medical abortion from about Ksh. 10,000 to about Ksh. 2,500 by Connecting them to consistent medical supplies, getting the supplies at wholesale price.

###### R (KII-Supplier 1)

Ma-Kare Combipack-Ksh. 500, Misoclear-450 ksh., MVA kit -4000 ksh.

###### R (KII-Supplier 2)

MA drugs are a bit expensive hence low demand.

###### R (KII-Pharmacist Trans Nzoiya)

The cost is affordable. Sometimes the drugs are supplied to us freely. Personally, I was given 10 packs of combipack.

## 4.0 DISCUSSION, RECOMMENDATIONS, AND CONCLUSIONS

### 4.1 Discussion

The findings indicated that most of the users of MA were under 25 years which is consistent with the findings by [11] in Tanzania that most young people prefer MA services compared to Manual Vacuum Aspiration (MVA). This distribution may be attributed to the fact that those seeking medical abortion were mainly youths who are either still in secondary schools, college, and Universities or just entering the labor market. The finding implies that the young women and girls are the main users of MA services.

Our results further indicated that a majority (92%) of the respondents reported that they were not aware of safe abortion practices. This result does not correlate with the findings of [3], who found that most clients seeking MA services in Kenya are aware of the existence of the MA services. The inadequate knowledge of safe abortion services points to a serious need to improve individual exposure to trusted information sources build urgency to support decision-making and prompt action. Data suggests a close association with geographic inequalities, cost, cultural, religious factors, and policy bottlenecks either at the national or county level which may be associated with the fact that abortion is highly stigmatized.

Further analysis indicate that many women and girls have reported using MA in the comfort of their homes. Women managing abortion on their own require sufficient information to help guide the process and the knowledge of where to access emergency care in case there is any complication. This finding is allude to the fact that self-medication is enshrined in many countries’ treatment-seeking habits, including in Kenya and Uganda where medicine sellers are often the first point of contact for girls and women who have health-related needs [12]. The finding implies that some women and girls will undergo a medical abortion completely on their own, provided they can get accurate information from or someone who can offer social/emotional support [13].

Knowledge of the gestation period within which it is safe to use MA is also very low and this finding is contrary to the findings by [14] whose results show that most women seeking MA services are aware of the gestation upon which those drugs should be used. Thus, courting potential complications that may ensue whenever medical abortion drugs are taken anyhow, without due consideration of the gestation period (safe window) and for effectiveness. The matter is worsened by the acknowledgment from the pharmacists that the women and girls seeking MA services deliberately lie about their gestation period so as not to be denied the services and or to pay lower user fees and this may be the reason why most lives of women and girls using MA may be endangered.

Further, those reporting having experienced medical complications arising out of an abortion procedure by use of misoprostol are a considerable number which is consistent to the findings by [15] from Malawi that complication rates from misoprostol outweighs that of combi pack. While most are directly attributed to the gaps in providing the correct dose and correct route of administration, this could inform the need for continuous support supervision to the pharmacists for greater responsiveness to the user needs. The implication is that combi pack is much more effective with low failure rates compared to misoprostol which most pharmacists do not have adequate knowledge in giving the correct dose as it depends on the gestation period.

A significant proportion of women and girls reported not being told anything about post-abortion contraceptives despite that it is particularly important in preventing unwanted and mistimed pregnancies for women between the reproductive ages (of 15-49 years). This finding contradicts the results from a study by [16] in Nigeria whose findings indicated that most women and girls were normally being informed of the existing contraception services whenever they visited the public health hospitals. This may be attributed by the observed existence of an information and awareness gap on matters of sexual and Reproductive Health that needs to be overcome. The implication is that this can lead to an unmet need for contraception, especially when the information provided does not support informed decision-making. Moreover, key dimensions of Sexual and Reproductive Health care access need to be analyzed to bring out key bottlenecks, especially for Adolescents and young people who seem to be the majority accessing MA services.

Nearly half (47%) of the women and girls reported being given post-abortion contraceptives they never wanted. The findings is concurrent to the findings by [17] in Zambia whose findings indicated that most pharmacists only sells the family planning methods they usually have in stock for profit making and not what clients’ needs. This points to a serious unmet need for contraceptives. Further, many who perhaps react to some types may be forced to use them since they are the only ones available even if they have some misgivings about them.

More than half (60%) of the women and girls mentioned that the pharmacist’s personnel were unable to address any fears and concerns they had regarding MA and contraceptives given. The finding agreed to the results from a study by [18] whose findings showed that pharmacists rarely offer comprehensive package of care including counselling to the women and girls seeking contraception services due to their limit in contraception service provision. This may be due to the counselling services offered by the pharmacists which is skewed to the contraception services they offer. The implication of this finding is that it can be a great bottleneck in the utilization of MA as the users will not feel safe and confident approaching the facilities. It could also point to palpable capacity gaps on the part of pharmacy personnel. Therefore, more hazardous abortions could be on the increase.

MA costs seem to vary greatly by inter and intraregional. Some are charged higher, some moderately, and some a bit low as below Ksh. 700 or offered services for free. This may be associated by the fact Ipas gave some seedstock to the pharmacists and that the providers deemed it wise to use such stocks to provide MA services to women and girls who might not have afforded them. There’s a need for standardization of costs otherwise prohibitive costs can begin to hinder access and utilization of MA or the personnel of pharmacists and clinics may turn the whole affair into a cash cow thus negating the whole concept of safe abortions (MA).

### 4.2 Recommendations

We have delineated key areas to inform on the MASU project scale-up. These have been presented as follows:

a. ***Access to:***
  i. Comprehensive adolescent and youth reproductive health (AYRH) - sexuality education, and contraceptive use education.
  ii. Post-abortion contraceptives. Women and girls of reproductive ages require this to prevent unintended (unwanted and mistimed) pregnancies. Without post-abortion contraceptives, unintended pregnancies could be on the rise, leading to increased demand for abortion services.
  iii. An array of contraceptives is preferred. Nearly 50% reported being forced to take contraceptives which are never their preference.
  iv. Affordable Medical Abortion drugs. Some users reported spending up to Ksh. 13,000 for medical abortion. The cost differentials among pharmacists need to be standardized.
b. ***Awareness Campaigns targeting:***
  i. Knowledge of safe abortion services among young women and girls. There is a need for urgent awareness campaigns. This was reported to be very low.
  ii. Information gaps on the gestation period within which one can safely use medical abortion pills still exist which exposes young women and girls to potentially serious complications.
  iii. The specific location of the pharmacists and clinics offering MA needs to be voiced out too.
  iv. Types of abortion pills. Clear information should be given on the types, side effects (if any), efficacy, and the convenient time to use them. Many reported they are not aware of the types of pills given.
c. ***Training and capacity building:***
  i. Pharmacy personnel needs further training on soft skills-handling abortion seekers, effective consultations that don’t make the users uncomfortable, confidentiality, follow-up mechanisms, and how to build stronger client-provider interrelationships.
  ii. Youth champions also need further training on how to conduct effective awareness campaigns in the counties of operation. Issues such as best approaches to women and girls, effective communication of matters around safe abortion, and how to forge stronger and healthier interrelationships with the users and pharmacists need to be targeted.
d. ***Drug suppliers:***
  i. Pharmacists embrace strategic purchasing from drug suppliers. This will involve buying abortion pills, contraceptives, and other related health consumables based on demand and efficacy. Less demanded pills need not form part of the expenditure.

### 4.3 Conclusions

The IPAS MASU project intervention, in the five counties of Western Kenya namely Busia, Siaya, Vihiga, Kisumu, and Trans Nzoia, has increased access to safe Medical Abortion self-use, enhanced availability of MA drugs in pharmacies, the improved service delivery of MA services through regular training of pharmacists. Further, the project has enhanced awareness about MA services among young girls and women through trained Youth Champions and pharmacists. In conclusion, the MASU project has significantly reduced cases of unsafe abortions and by extension deaths and medical complications associated with them. To realize more gains, the project needs to be scaled up within the five counties and beyond, specifically to target the rural areas where cases of unsafe abortion are still thought to be rampant.

## Data Availability

Data is available within the organization's cloud server and can be shared upon requests

## 5.0 ACKNOWLEDGEMENTS

The authors are grateful to the consultant, Dr. Francis Omondi of Kenyatta University who worked with Ipas team to complete this process evaluation. We also want to specifically thank Japheth Ogol for his immense contribution in this evaluation including development of Terms of Reference for the Consultants, Developing and digitalization of the data collection tools, validation of the analysis, and writing of the manuscript. In addition, we are grateful to all pharmacists and private clinics in Kenya and MA users for their involvement in this process evaluation survey. We are also saluting the Ethical Review Committee (ERC) of Jaramogi Oginga Odinga Teaching and Referral Hospital (JOOTRH) for approving the research through a permit Ref. IERC/JOOTRH/464/21 to carry out the study. We also appreciate Ipas Africa Alliance implementing team of Kenya for their large input in initiating and coordinating this research project. We also appreciated all the authors who contributed to reading and approving the final manuscript.

